# X-COVNet: Externally Validated Model for Computer-Aided Diagnosis of Pneumonia-Like Lung Diseases in Chest X-Rays Based on Deep Transfer Learning

**DOI:** 10.1101/2024.05.20.24307627

**Authors:** Jorge Félix Martínez Pazos, Jorge Gulín Gonzales, David Batard Lorenzo, Arturo Orellana García

## Abstract

Since the appearance of COVID-19, the accurate diagnosis of pneumonia-type lung diseases by chest radiographs has been a challenging task for experts, mainly due to the similarity of patterns between COVID-19 and viral or bacterial pneumonia. To address this challenge, a model for the computer-aided diagnosis of chest X-Rays has been developed in this research. This model might contribute to substantially increasing the accuracy of the diagnosis. This approach is based on supervised learning using neural networks, where the quality of the result depends on the quality of the dataset used during training. Image data augmentation techniques, hyperparameter adjustments and dropout layer contributed to achieve high performance values on test data in multi-class classification. The experiments conducted to evaluate the model yielded that it detects and classifies domain classes with an accuracy of 99.45% on training data, 99.27% on validation data and 99.06% on selected test data. The main contribution of this paper is X-COVNet a new Deep Convolutional Neural Network model using Deep Transfer Learning through the Xception architecture for the assisted diagnosis of COVID-19, pneumonia or healthy patients, trained on COVID-19 Chest X-Ray Database and evaluated through two external databases, which give the model novelty within the lack of external validation in all the literature reviewed.

## 1. Introduction

During the COVID-19 pandemic crisis, plenty of health institutions suffered periods of collapse and their workers had to deal with the burden of attending to each of the patients. Although other techniques were later used to diagnose the disease, resorting to a chest X-ray is the most common when a patient has signs of respiratory disease such as COVID-19 or pneumonia. In 2017, more than 808 000 children under five years of age deceased due to pneumonia, accounting for 15% of all deaths in children under five years of age. Individuals who are susceptible to developing risk pneumonia encompass those who are above the age of 65 as well as those with underlying medical conditions [1]. Currently, it is possible to accurately diagnose pneumonia. However, it is a difficult task, requiring the review of a chest X-ray which must be performed by highly trained specialists. This disease usually manifests as an area of increased opacity on the chest X-ray [2]. However, the diagnosis of pneumonia from X-rays is made difficult in early 2019 by the similarity of patterns between pneumonia and COVID-19. Differences between pneumonia and COVID-19 in X-rays are difficult for the human eye to perceive, but intelligent system-assisted diagnosis could result in greater accuracy. For example, models based on Artificial Intelligent (AI) can study the internal patterns in the pixel array of the image. Specialists are often faced with reading large volumes of images during each work shift. Hence, having a tool for computer-aided diagnosis might positively impact the efficiency and effectiveness of the outcome.

Intelligent systems based on deep learning have been used in multiple solutions for medicine and healthcare. Yu et al. [3], show a non-exhaustive list of the potential of AI applied in medicine grouped into, basic biomedical research, translational research and clinical practice.

The field of medical imaging for computer-aided diagnosis has been covered by several authors with excellent results, studies discussed below perform the classification of chest X-rays. Hashmi et al. [4], proposed to obtain the optimal weights for five deep convolutional neural network architectures - ResNet18, DenseNet121, Inception, Xception and MobileNetV2 - and feed the weighted predictions into a weighted classifier module to obtain the final weighted prediction to identify healthy patients or patients with pneumonia. Chouhan et al. [5] used an ensemble model consisting of multiple pre-trained deep neural network models. Toğaçar et al. [6], combined features from different deep learning models. Ayan et al. [7], applied transfer learning and fine tuning to VGG16 and Xception architectures. Civit-Masot et al. [8] also applied transfer learning to the VGG16 architecture to distinguish between COVID-19, pneumonia and normal classes. Owida et al. [9] proposed a method for extracting effective features from chest X-ray images using wavelet analysis and the Mel Frequency Cepstral Coefficients (MFCC) method. These features were then used in a classification process using a Support Vector Machine (SVM) classifier. In a study by Lee & Lim [10], fine-tuning techniques were applied to the DenseNet201 architecture to improve its performance in detecting COVID-19 cases from chest X-rays. Jyoti et al. [11], present a new approach to decompose chest radiographs from two different datasets using a two-dimensional (2D) tunable Q-wavelet transform (TQWT) based on a memristive crossbar array (MCA). The decomposed images were then classified as either COVID-19 or non-COVID-19 using convolutional neural network (CNN) models, specifically ResNet50 and AlexNet. Their results were achieved with less complexity, energy consumption and performance compared to conventional techniques. Dalvi et al. [12] proposed to apply transfer learning and tuning over DenseNet-169 architecture, where the data preprocessing is performed using the Nearest-Neighbors interpolation technique.

Considering the significant benefits of Intelligent systems based on deep learning, this research aims to create a predictive model based on deep learning techniques. This model enables computer-aided diagnosis of respiratory diseases in chest radiographs using a multiclass classification between normal/healthy, pneumonia and COVID-19. This approach is based on supervised learning using neural networks where the quality of the result depends on the quality of the dataset used during training. Image data augmentation techniques, hyperparameter adjustments and dropout layer contributed to achieving high performance values on test data in multi-class classification.

The following is an outline of the contributions which have been provided in order to draw attention to the relevance of the work that will be presented by this study:

- X-COVNet is the main contribution of this proposal, a Deep Convolutional Neural Network model using Deep Transfer Learning through the Xception architecture for the assisted diagnosis of COVID-19, pneumonia or healthy patients, trained on COVID-19 Chest X-Ray Database and evaluated through two external databases that give the model novelty within the lack of external validation in all the literature reviewed.
- X-COVNets an open-source GitHub repository aiming at a medical imaging model library. Available online under MIT License: https://github.com/WiseGeorge/X-COVNets.
- The use of transfer learning to achieve better results in less time and at less cost.
- Image data augmentation techniques are applied and their potential to significantly improve the results of the image classification task is demonstrated.
- The results confirmed the reliability of the proposed model in multi-class classification, obtaining remarkable values of around 99% and 95% for internal and external evaluation, respectively, for each metric assessed.

The rest of the paper is structured as follows. In Section 2, the main materials and methods adopted to develop and evaluate the proposal are described. The results of the developed model are analyzed in Section 3. The comparison of the results presented in this research with those reported in the literature is discussed in Section 4. Finally, conclusions and future work are presented.

## 2. Materials and Methods

### 2.1 Dataset

The COVID-19 Chest X-Ray Database, which was officially published on Kaggle and developed by a team of researchers from Qatar University in Doha, Qatar and Dhaka University in Bangladesh, along with their associates from Pakistan and Malaysia in conjunction with medical professionals, was deemed the most affordable option for the proposed approach [13]. The database contains chest X-ray images for healthy, COVID-19 positive and Normal and Viral Pneumonia cases. The suitability of this database is given by the fact that the data were collected from various publicly available datasets, online sources, and published articles, and include images from several regions, including the Americas and Europe, since images were sourced from several reputable institutions, such as the Radiological Society of North America, the Institute for Diagnostic and Interventional Radiology at Hannover Medical School in Hannover, Germany, the Italian Society of Medical and Interventional Radiology (SIRM), and the Medical Imaging Databank of the Valencia Region (BIMCV). The inclusion of images from these different sources ensures that the training dataset is representative of a wide range of radiological data.

The database contains 2500 images of the normal class, 1345 of pneumonia, and 3616 of COVID-19. Originally, the database had more than 10,000 images belonging to the normal class, to balance the dataset, a random value between the maximum and minimum of the other classes was selected, resulting in 2500 images for the normal class. Figure 1 shows the first contact with the database.

**Fig. 1:**
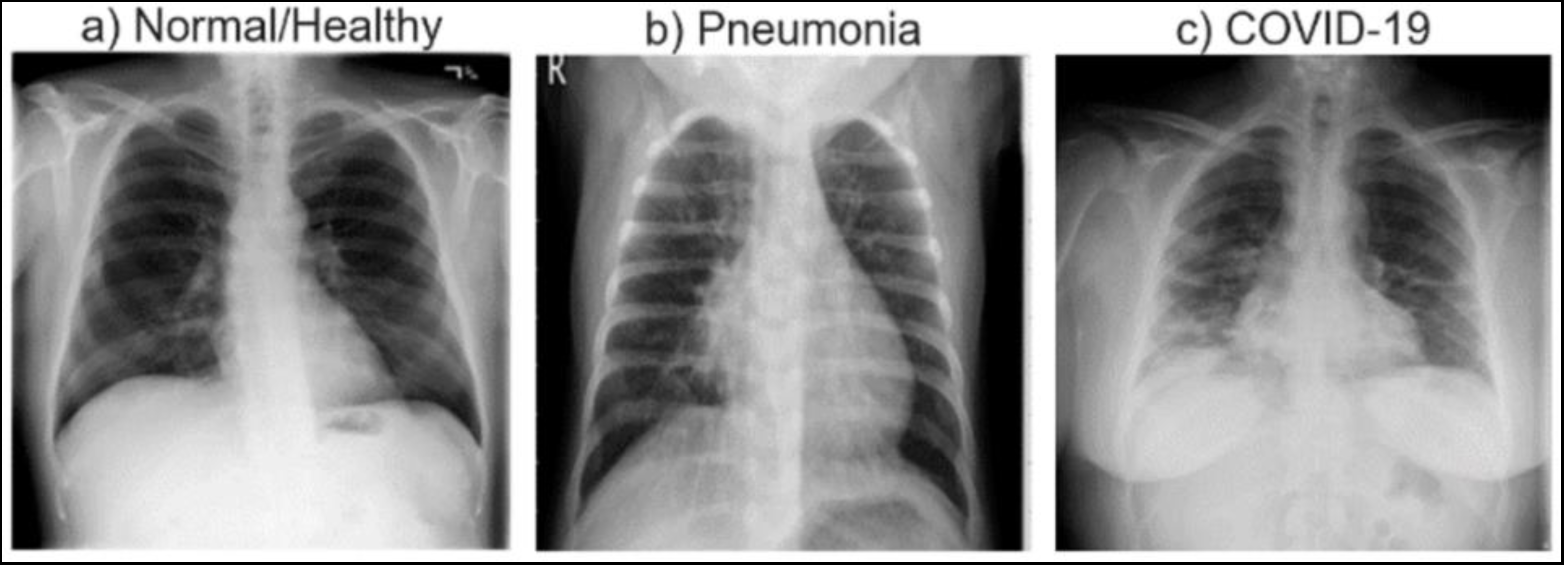
Chest radiographs of a) a healthy individual, b) an individual with pneumonia, and c) an individual with COVID-19.

### 2.2 Performance Metrics

All performance metrics used in this paper are discussed below. In the definitions and equations mentioned below, when classifying normal/healthy, pneumonia and COVID-19 in patients being category_var any value of the domain to be classified; true positive (TP) denotes the number of category_var images identified as category_var, true negative (TN) denotes the number of ¬category_var images identified as ¬category_var, false positive (FP) denotes the number of ¬category_var images incorrectly identified as category_var images and false negative (FN) denotes the number of category_var images incorrectly identified as ¬category_var [4].

- **Accuracy:** Indicates how close the measured value is to a known value.

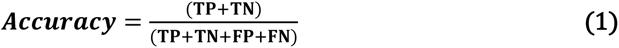

- **Precision:** Indicates how accurate the model is in terms of those predicted to be positive.

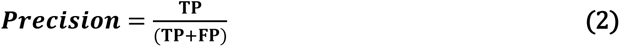

- **Recall:** Calculates the number of real positives that the model was able to capture after labeling it as positive.

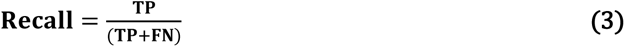

- **F1:** Provides a balance between precision and recall.

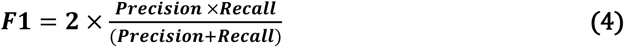

- **Confusion Matrix:** Summarizes the classification performance of a classifier [14].
- **ROC (Receiver Operating Characteristics) Curve:** Graphical representation of the relationship between a classifier’s sensitivity (true positive rate) and 1 - its specificity (false positive rate).
- **AUC (Area Under the Curve) Score:** Measure of the degree to which a classifier can distinguish between classes. A higher AUC score indicates a better ability to differentiate between classes.

### 2.3 Transfer Learning & Xception Architecture

Transfer learning is a technique that involves enhancing the learning process for a new task by transferring knowledge from a related task that has already been learned [15]. The use of pre-trained models as an initial foundation for tasks related to computer vision and natural language processing is a prevalent methodology within the field of deep learning [16]. Xception is a deep convolutional neural network architecture that employs depth-wise separable convolutions. The author postulated that inception modules within convolutional neural networks serve as a transitional phase between standard convolution and depth-wise separable convolutional operations [17].

### 2.4 Developed Predictive Model based on Deep Learning Techniques

The workflow of the proposed model is shown in Figure 2. The first step is the acquisition of knowledge about the domain of the subject to be worked on. In this case, medical imaging, the techniques that are used at a professional level and the standards that govern it. The in-depth study of the standard DICOM medical imaging format allows for a greater scope of the proposed model. After data acquisition, exploratory data analysis is used to obtain insights from the dataset. The architecture of the deep learning model is defined by 3 blocks data preprocessing - data augmentation, model building - fine tuning and callbacks. The preprocessing is worked exhaustively because it has a direct impact on the model’s performance, model building - fine tuning block and callbacks block are inter-connected due to the presence of a feedback mechanism between them.

**Fig. 2:**
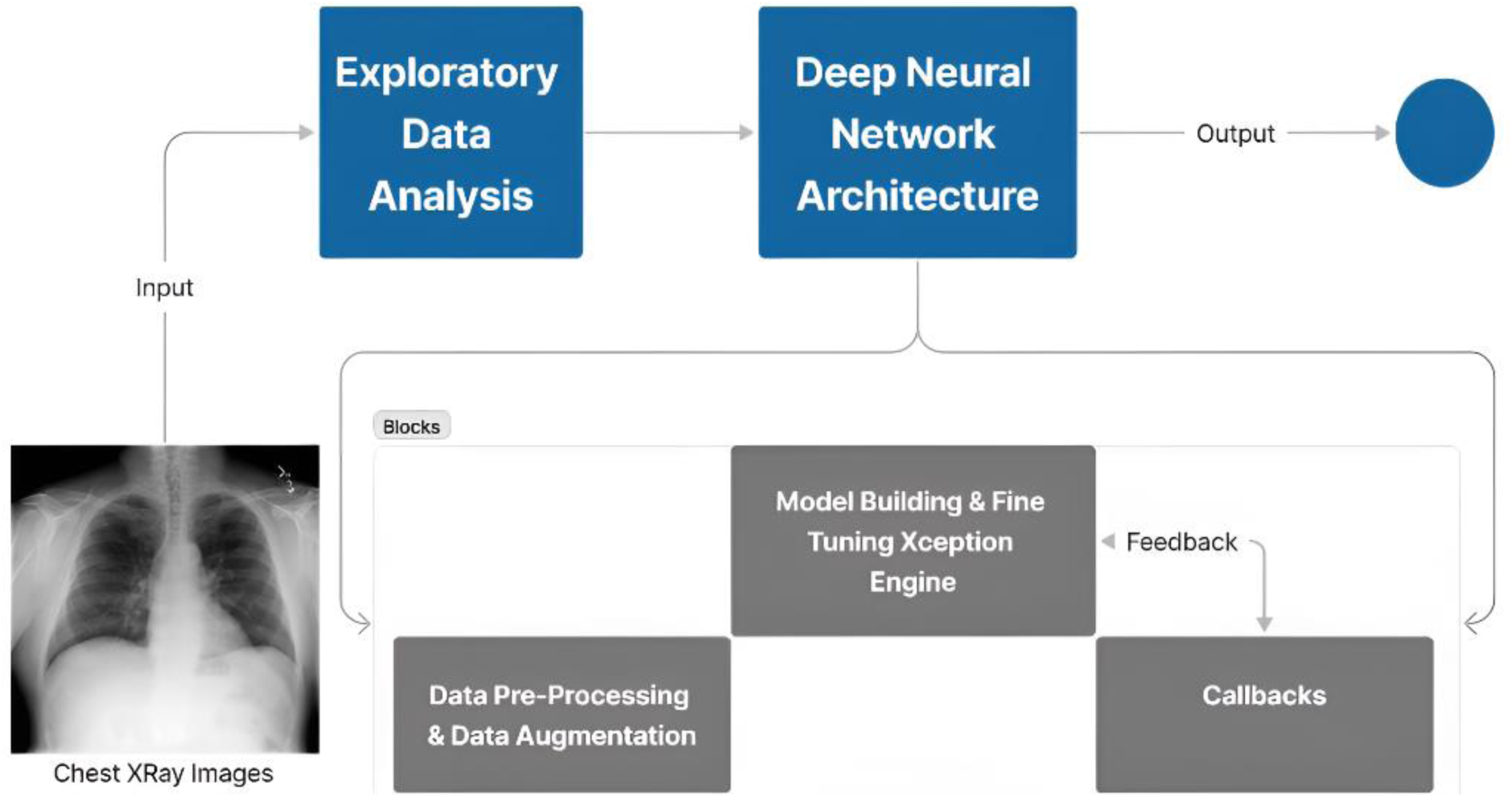
Workflow diagram of the proposed model.

With new advances in artificial intelligence, a range of tools and libraries have become popular that make it easier to build neural network models. The proposed approach was developed following the coding standard for the Python programming language, PEP8. Likewise, the proposal uses the main libraries for machine learning, deep learning and data science: Tensorflow 2.10.0 and ScikitLearn 1.2.1 for modeling; Numpy 1.23.3 and Pandas 1.4.4 for the analysis, study and manipulation of the dataset; Matplotlib 3.5.3 and Seaborn 0.12.0 for visualizations; and Keras Tuner 1.1.3 for hyperparameters fine tuning. To achieve improved optimization and more efficient handling of uncertainty, convolutional neural networks and transfer learning techniques were employed. These methods have demonstrated exceptional performance in computer vision tasks [18]. In the following sections, each component of this model is described.

### 2.5 Exploratory Data Analysis

During the data reading process, the images pixel arrays and their respective classification were archived in a dataframe. Pandas dataframe and numpy array are the data types to be worked on. These allow adequate data manipulation and more efficient and faster processing. The images were formatted to RGB with a size of 224x224 for optimal processing of the predictive model. Working with high resolution images would lead to a high level of processing and time cost. As the scatter plot depicted in figure 3 shows, there is no significant dispersion of values concerning the total, 91.6% of the images have values distributed in (75 ≤ x ≤ 175) for mean and (30 ≤ y ≤ 80) for standard deviation. Images with values outside this range have very specific characteristics that can provide the predictive model with patterns that are not common in chest radiographs. Hence, this small number of outliers are not treated in this research.

**Fig. 3:**
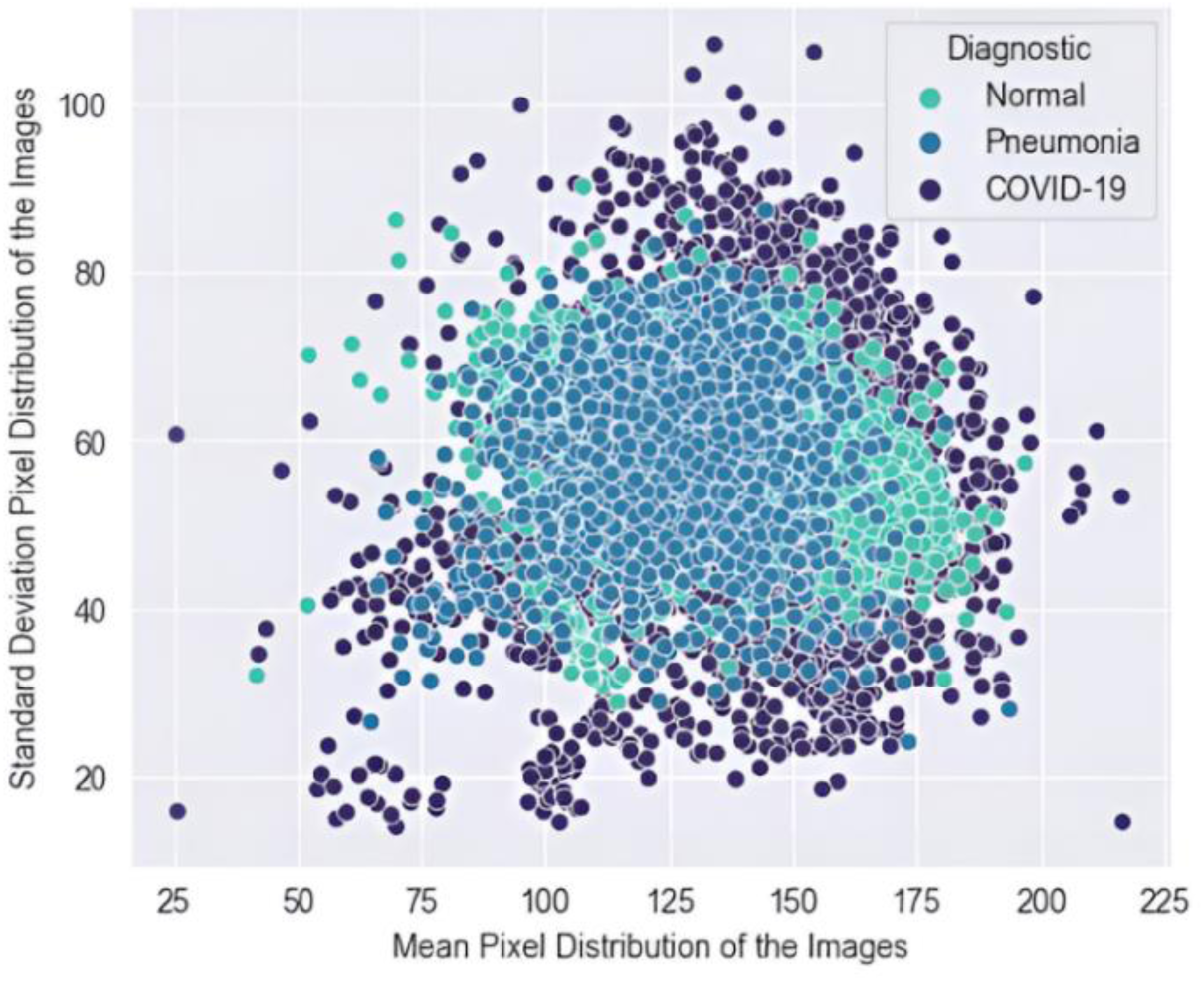
Scatter plot showing the mean and standard deviation of pixel values in images.

Data preprocessing, normalization, and correction were rigorously performed to mitigate potential biases in the predictive model, enhance its generalizability, and prevent the occurrence of false positives or false negatives. A detailed manual analysis of stochastic image sets was conducted to ascertain the number of radiographs with COVID-19 or pneumonia classification among patients admitted with visible medical wiring, which could lead to erroneous learning and misclassifications by the model, based on the assumption that all patients exhibiting these features belong to a given classification. Images with these features are sparsely represented in the dataset, obviating the need for their removal or treatment.

### 2.6 Deep Neural Network Architecture

#### 2.6.1 Data Preprocessing & Data Augmentation

The dataset was split into three portions, 80% of the images were taken for the training set; 20% were taken for the validation set, from this 50% were taken for the test set. As a result, the final distribution of the data set was 80% for training, 10% for validation, and 10% for testing. Table 1 show the dataset’s final distribution.

**Table 1.** Distribution of train, validation and test set.

| Set | No. Images |
| --- | --- |
| Train | 5968 |
| Validation | 747 |
| Test | 746 |

The images from the training and validation set which had already been formatted to a scale of 224x224x3 were normalized. The pixel values were taken from the range 0-255 to 0-1 for optimal computational processing. Previously 71x71x3 and 100x100x3 were used, but tests with these image formats resulted in low performance levels compared to the applied one. Keeping the images with a resolution around 200 pixels and in RGB scale translates into more patterns in the pixel array that can be studied by the model without increasing the processing time too much. To increase the diversity of the dataset and improve model performance, various image data augmentation techniques with a probability of 100% such as rotation with 25 degrade limit, horizontal flipping, random brightness adjustment, and RGB shifting are applied using Albumentations library, as depicted in figure 4 [19, 20].

**Fig. 4:**
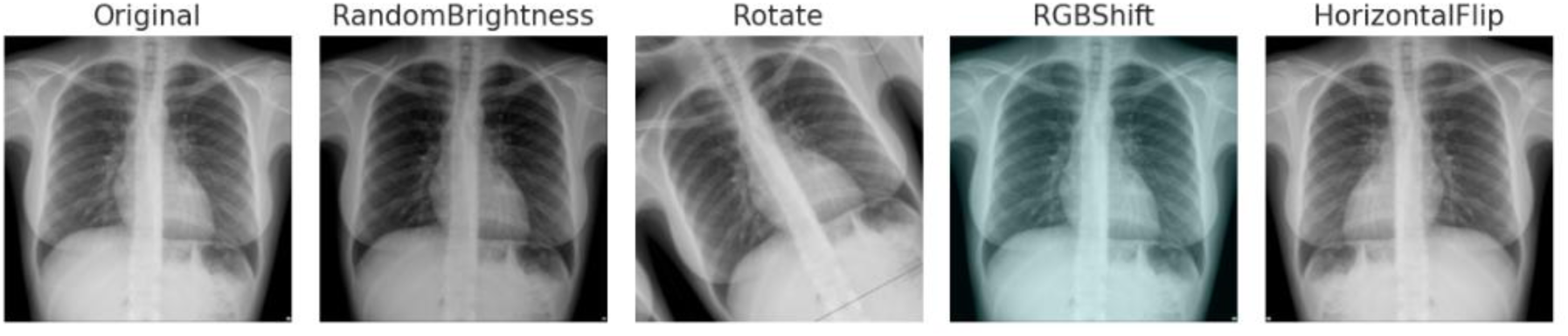
Representation of the applied image data augmentation techniques.

#### 2.6.2 Model Building & Fine Tuning Xception Engine

The proposed model is initialized and configured using the Xception architecture and adding new configurations, input layer 224x224x3, pretrained architecture weights were frozen and used the ’imagenet’ weights which are pretrained on ImageNet database organized according to WordNet hierarchy [21]. In the output of the Xception architecture, was added a 2D Global Average Pooling layer. The parameter compression ratio is exponentially high in this type of layer resulting in a 2D dimensionality of the form (batch_dimension, n_channels). This is different to the Flatten layer commonly used to feed fully connected layers, which only restructures the matrix to a single dimension [22]. Followed by a Dropout layer with 75% probability to avoid possible overfitting mainly due to the extension of the pretrained model, and a Batch Normalization layer to normalize the inputs. This layer applies transformations that keep the mean output close to 0 and the standard deviation output close to 1 [23, 24]. It is concluded with a Dense output layer with 3 neurons referring to the three classes in the domain, using the softmax activation function. Figure 5 shows the input and output flow of the deep convolutional neural network with the proposed configurations, where the red squares represent the main modifications described above.

**Fig. 5:**
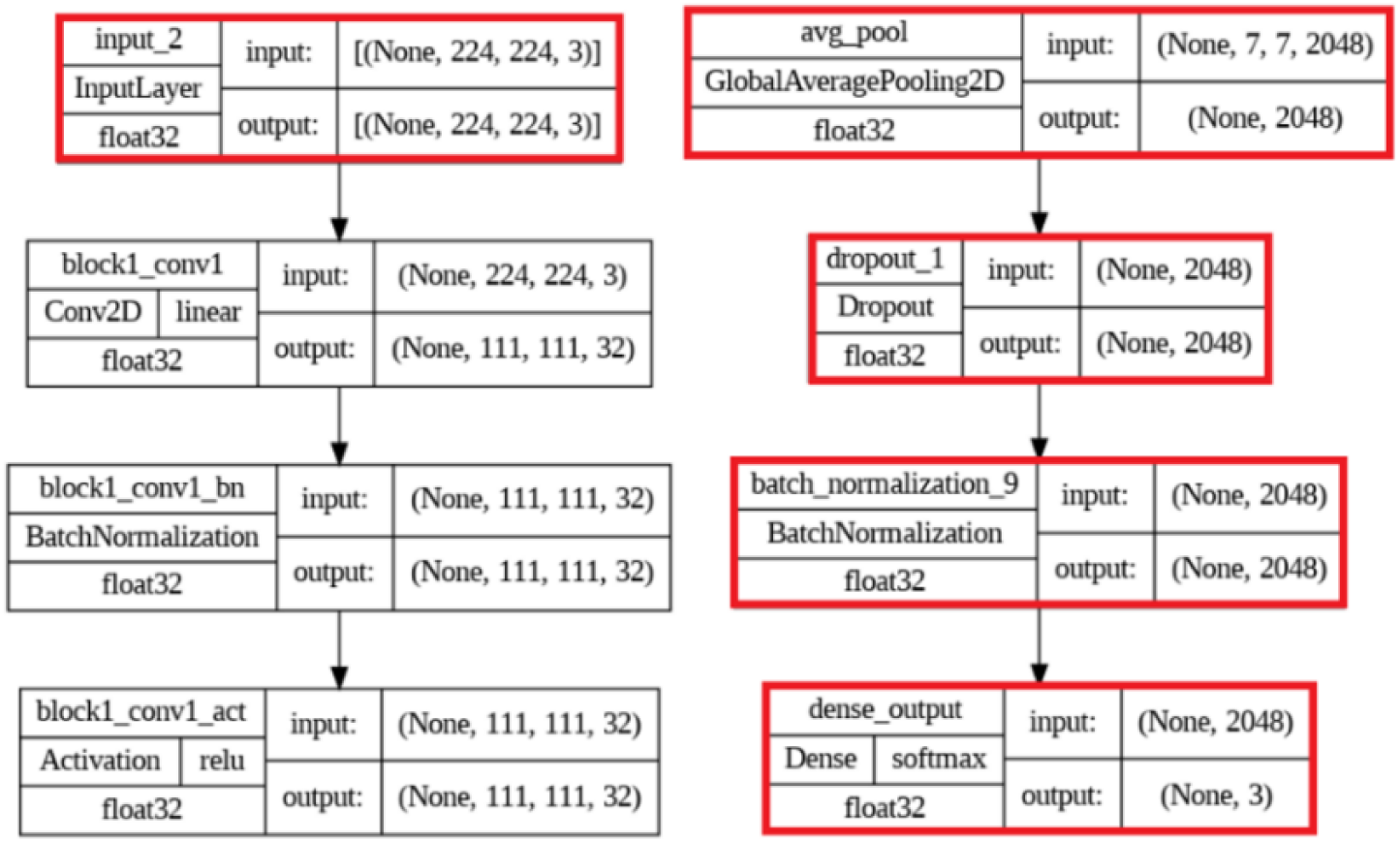
Input and output flow of the proposed model.

The model is compiled using the Adaptive Moment Estimation (Adam) optimizer with a learning rate of 3e-4, the loss function categorical crossentropy, the metric accuracy and a batch size of 32. The values corresponding to learning rate and batch size were obtained from the fine tune process using keras tuner, the hyperparameters search was run on [0.03, 0.003 0.0003] and [16, 32, 64] for learning rate and batch size respectively [25]. The recommended values for the batch size parameter in low-performance computing were selected. The learning rate values were chosen because, although the default value for the Adam optimizer is 0.001, a widely used value is 0.0003 [26]. Therefore, an interval from 0.03 to 0.0003 was used, decreasing by a factor of 10.

#### 2.6.3 Callbacks

Four callbacks were used during the training process, early stop, learning rate reduction, tensorboard and checkpoint [22]:

- **Early Stop:** Stops the training process after no better values are achieved in the loss metric.
- **Learning Rate Reduction:** Reduces the learning rate of the model, set at 3e-4 by monitoring the loss on validation data allowing for better optimization and accuracy of the model on validation data.
- **Tensorboard:** Tool included in the Tensorflow Framework for real-time visualization of all variables and the model’s behaviors, favoring the optimization of hyperparameters.
- **Checkpoint:** Allows to save the weights and biases of the model in a certain state as it is trained, in this case it saves each time the model improves depending on the metric val_loss.

The value of the training epochs during the first test was 35 epochs. After the first test training, the early stop callback stops the training process at epoch 15. The value of patience used in this callback was exceeded, 5 in this case, because the value of the metric val_loss did not improve from this epoch. Therefore, 15 is set as the definitive value for the training epochs. Table 2 summarizes the hyperparameter settings of the deep neural network.

**Table 2.** The hyperparameter settings of the deep neural network.

| Parameter | Value |
| --- | --- |
| Image size | $224 \times 224 \times 3$ |
| Batch size | 32 |
| Dropout rate | 0.75 |
| Training epochs | 15 |
| Optimizer | Adam |
| Learning rate | $3e-4$ |
| Loss Function | Categorical Crossentropy |

The techniques of image data augmentation, learning rate reduction, dropout layer together with real-time monitoring of hyperparameters through the tensorboard were specifically worked on to avoid overfitting of the proposed model. The model training is performed using an Intel(R) Core (TM) i5-7300HQ computer with 2.50GHz CPU, NVIDIA GeForce GTX 1050 2GB GPU, 8GB RAM.

## 3. Results

### 3.1 Model History

After training the model on the training set for 15 epochs an accuracy of 0.9945 on training data and 0.9927 on validation data was obtained. Figure 6 shows the behavior of the metrics during each epoch corresponding to loss, accuracy, validation loss, validation accuracy and learning rate reduction.

**Fig. 6:**
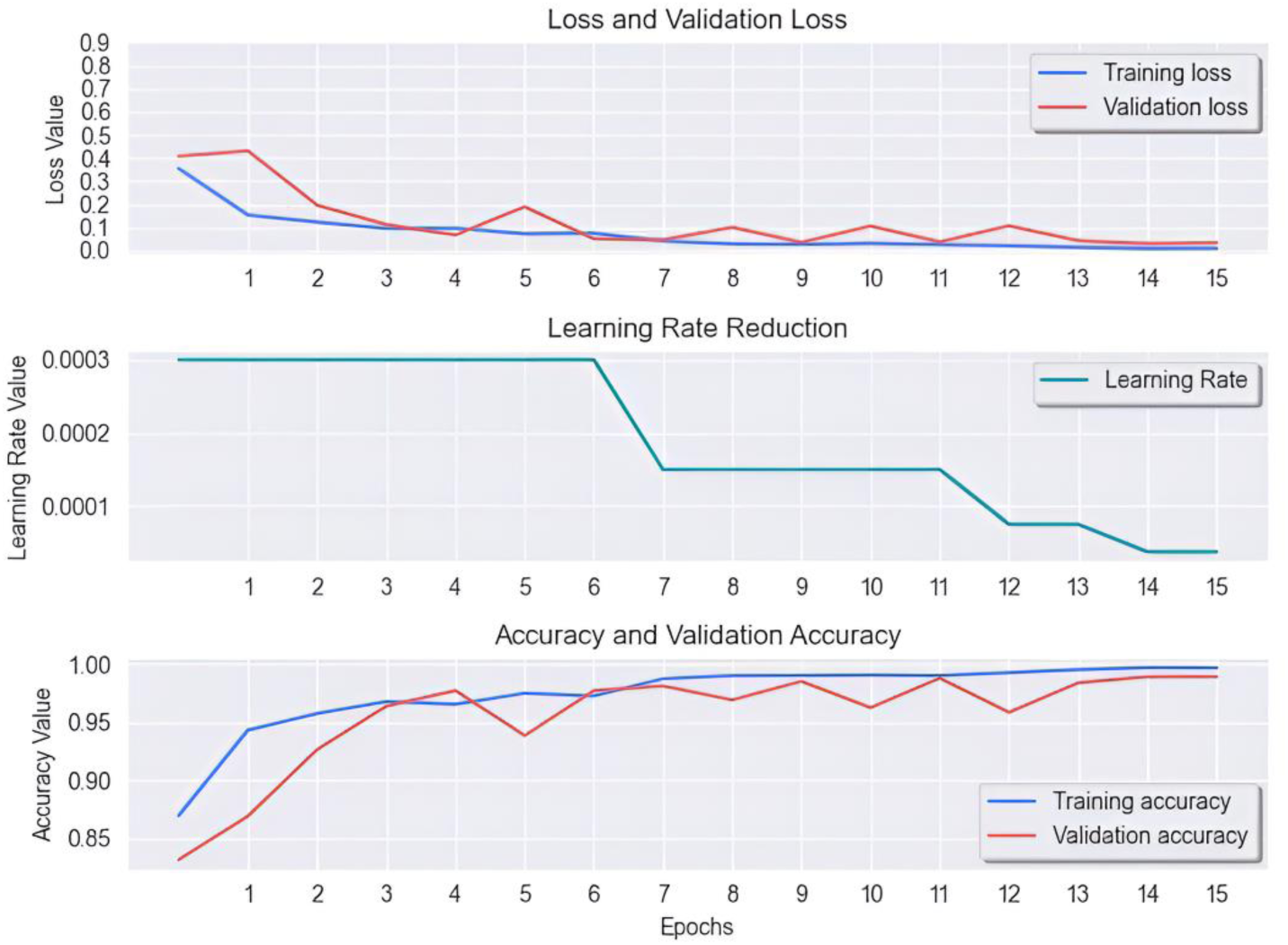
Graphical representation of the model’s training process.

The model exhibits robust generalization capabilities, as evidenced by its consistent performance across both the training and validation datasets. The benefits of transfer learning: a higher start, a higher slope and a higher asymptote are noticeable in the model training history as shown in figure 6 [27]. After concluding the training, it is visible the considerable contribution of the callback learning rate reduction to the performance of the model on validation data. Also shows how the reduction of the learning rate stabilizes the model keeping it close to the best metrics, the decay of the learning rate is also translated in a smaller amplitude between the training and validation curve. This is due to the fact that the lower the value of the learning rate used in the gradient descent implemented by the Adam optimizer, the smaller its displacement in search of the global minimum, therefore it can locate this or very close values with greater effectiveness, having less possibility of deviating to less favorable situations [20]. The best values for each metric are reached at epoch 15, so this is the saved configuration of the model via the checkpoint callback.

### 3.2 Assessing the Model Performance

The model was evaluated on the test set after completion of the training phase, model evaluation: 69s 3s/step - loss: 0.0393 - accuracy: 0.9906. Its performance was validated using accuracy, recall, precision, F1 and confusion matrix. The model achieved 100% recall for the COVID-19 class, indicating that all instances of this class were correctly identified; such high sensitivity in detecting patients with COVID-19 suggests that there were no false negatives for this class. Results detailed in table 3 indicate that the model performed slightly better on the COVID-19 class than on the normal and pneumonia classes, suggesting that the model is more effective in identifying patients with this disease.

**Table 3.**
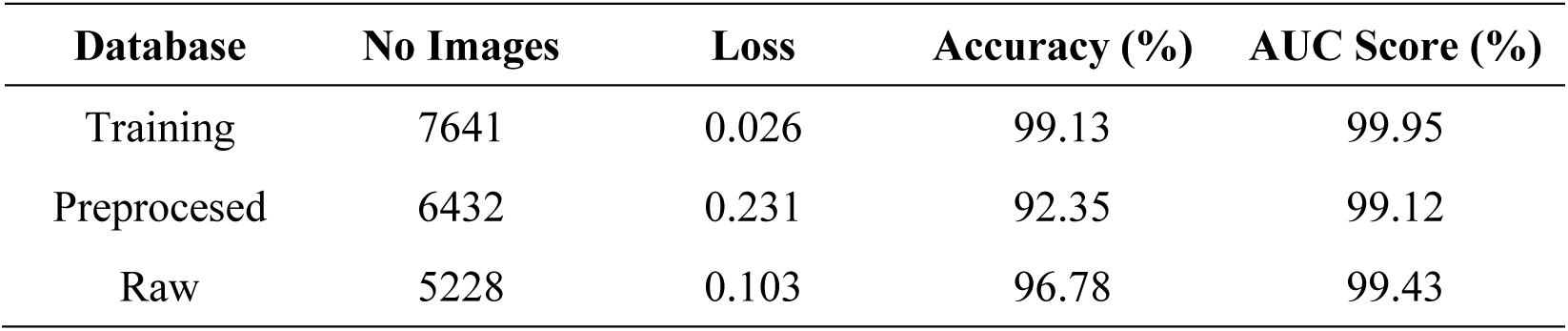
Metrics of the model evaluation on external databases.

The confusion matrix obtained from the evaluation of the model yields high performance values. The diagonal entries of the matrix correspond to the correctly predicted values, while the off-diagonal elements represent cases of incorrect predictions. As indicated in the figure 7, the model exhibits a remarkably low misclassification rate.

**Fig. 7:**
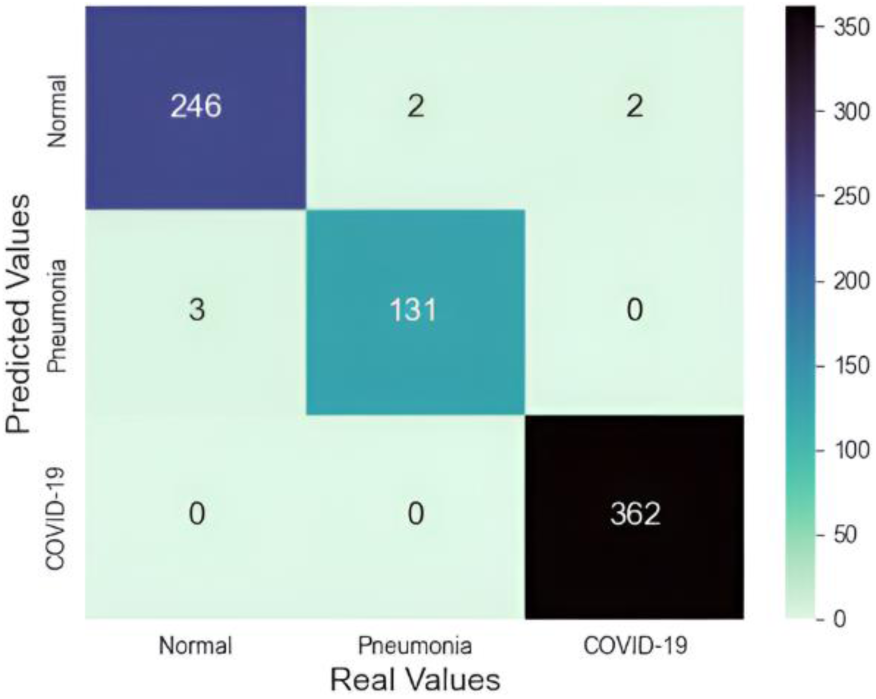
Confusion matrix for the test set of 746 images.

A set of images were randomly selected from the test set for classification using the proposed model. Figure 8 provides a visual representation of each image, accompanied by its respective predicted classification from the X-COVNet model, indicated by the label on the x-axis (Predicted Label: PL), and its actual classification, indicated by the label on the y-axis (True Label: TL). The previous results demonstrate the remarkable performance of the model in correctly classifying chest X-rays.

**Fig. 8:**
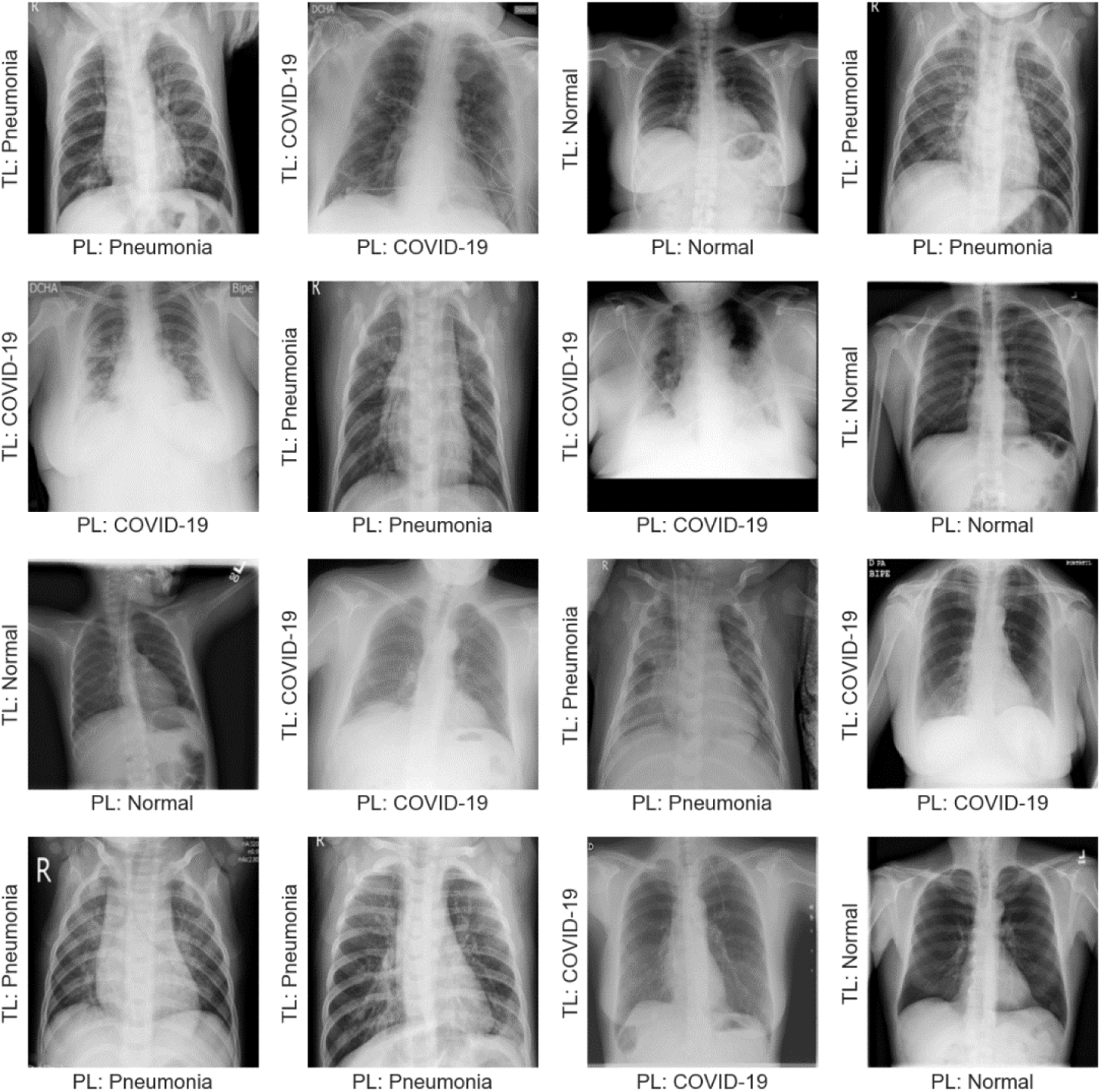
Classification results of chest radiographs using the proposed model.

### 3.3 Assessing Generalization Capabilities on Real-World Data

It is important to evaluate predictive models on unseen data, especially in the context of medical imaging. If a model is not thoroughly evaluated with data external to the training process, there is a risk that it will not perform as well in real-life scenarios as it did during training. This could lead to incorrect diagnoses and potentially harmful therapeutic decisions. By evaluating the model with new data, practitioners can gain a more accurate understanding of its real-world performance and increase confidence in its use for assisted diagnosis. This can help ensure that the model is reliable and safe for use in clinical settings.

This process involves the use of two external chest radiograph databases containing a large number of images of 6000 approximately. These databases allow for a comprehensive evaluation of the model’s performance on both raw and preprocessed images. The evaluation process begins with an assessment of the model on the training database COVID-19 Chest X-Ray Database [13]. This serves as a reference point for further evaluation on external databases. The first external database which will be called Raw Database, Chest X-ray (Covid-19 & Pneumonia), is derived from three previously published datasets and contains a total of 6432 images. One of these datasets is ieee8023 covid-chestxray-dataset, an official database approved by the University of Montreal’s Ethics Committee and developed by Joseph Paul Cohen and his team at Mila, University of Montreal [28]. This database contains raw images in various formats and sizes, including images with figures and other artifacts as detailed in figure 9.

**Fig. 9:**
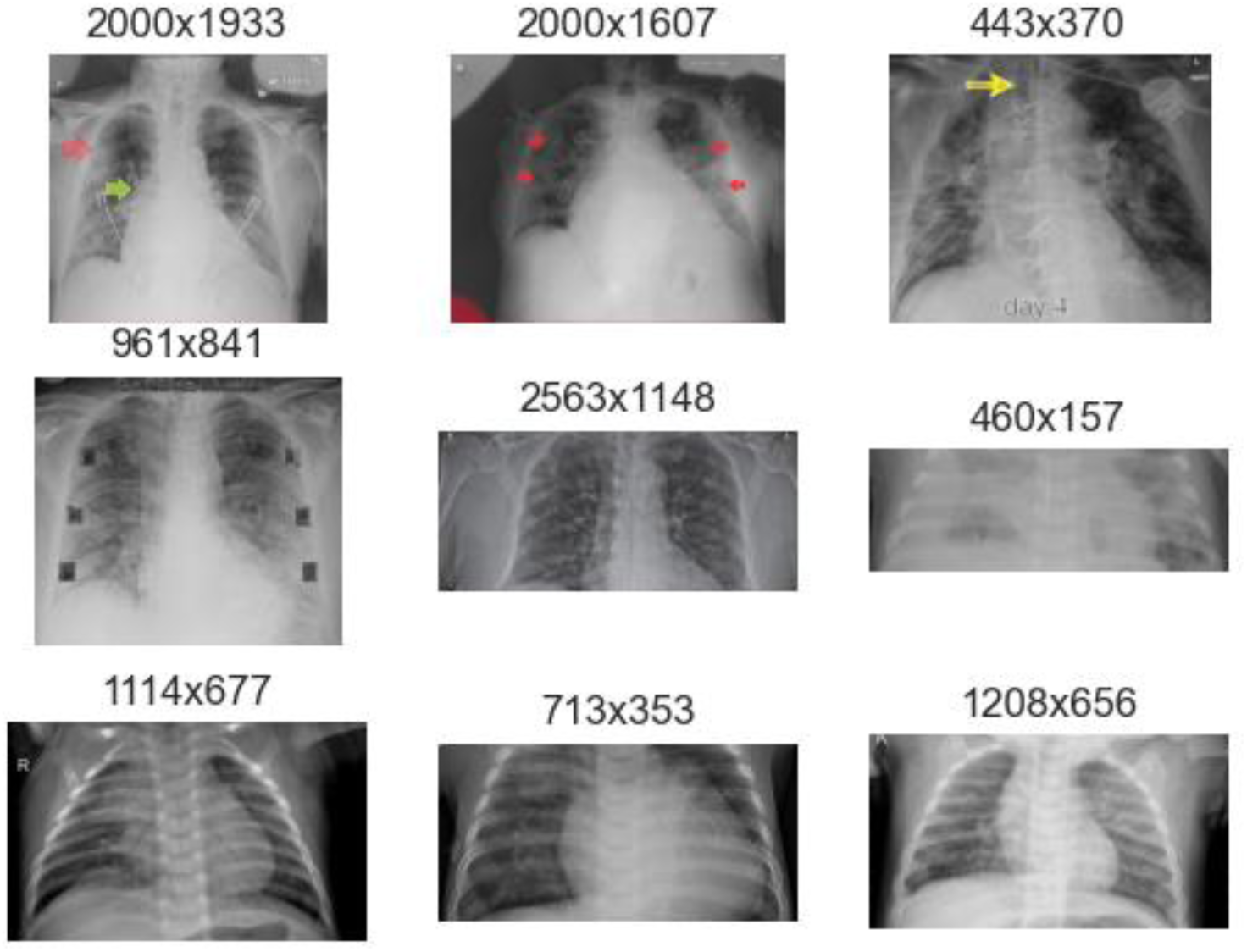
Sample images from the raw database.

The second external database used is the Chest X-Ray Images Database, called Preprocessed Database, which has a total of 5228 images and contains preprocessed images resized to 232x232 in png format, including an even larger number of images with artifacts such as pointers and numbers, as detailed in figure 10. It uses information from medical websites such as eurorad.org, radiopedia.org and coronacases.org [29].

**Fig. 10:**
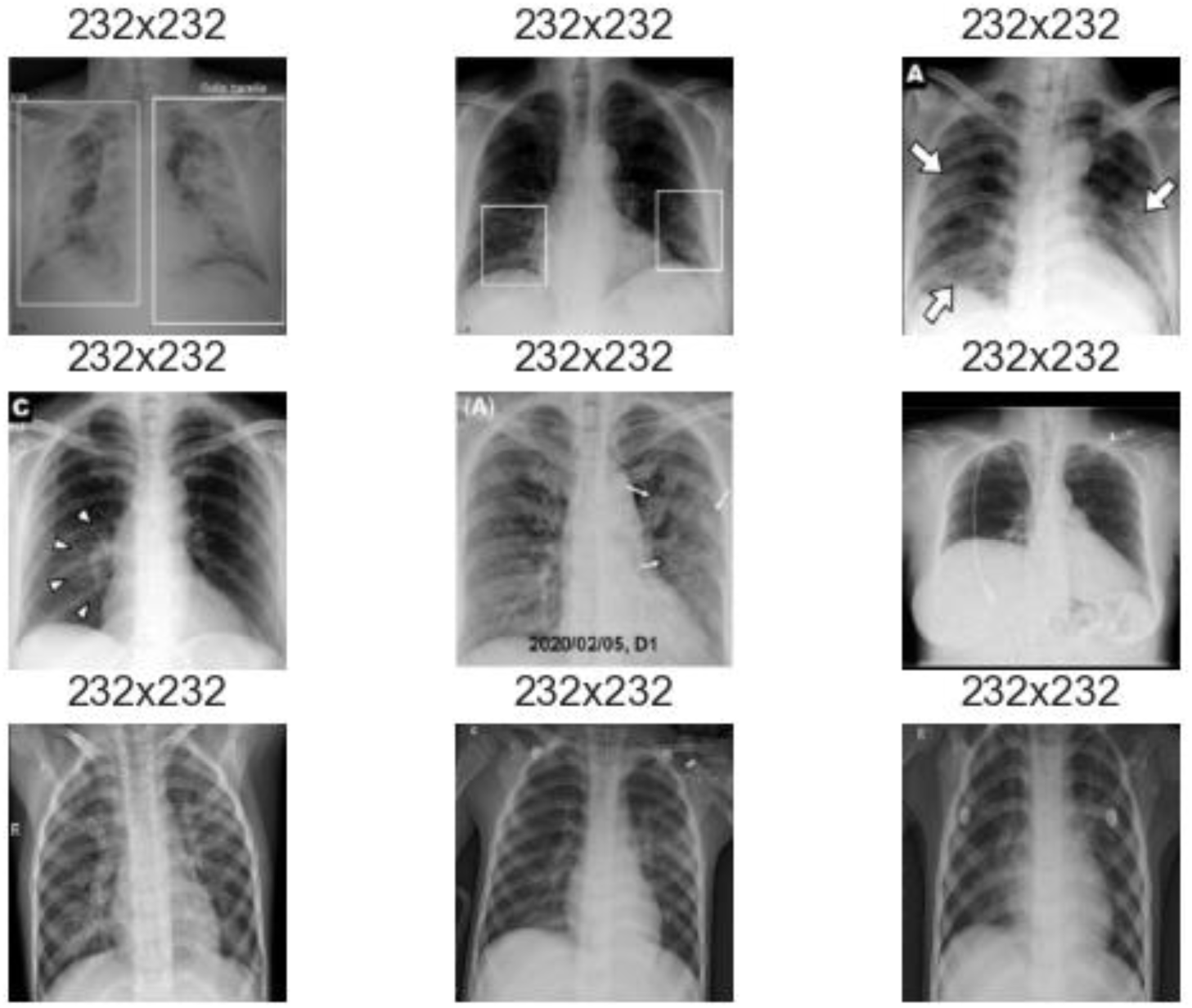
Sample images from the preprocessed database.

To gain a deeper understanding of the classification model on the three classes represented in the assessment databases, the graph in Figure 11 shows the ROC curve for One vs One and One vs Rest, as well as the AUC score. The AUC score is an indicator of the performance of the model by domain class, evaluating true positives vs false positives. The OvO and OvR strategies are used to adapt the ROC curve and AUC score metrics, originally intended for binary classification, to multi-class classification problems [30, 31].

**Fig. 11:**
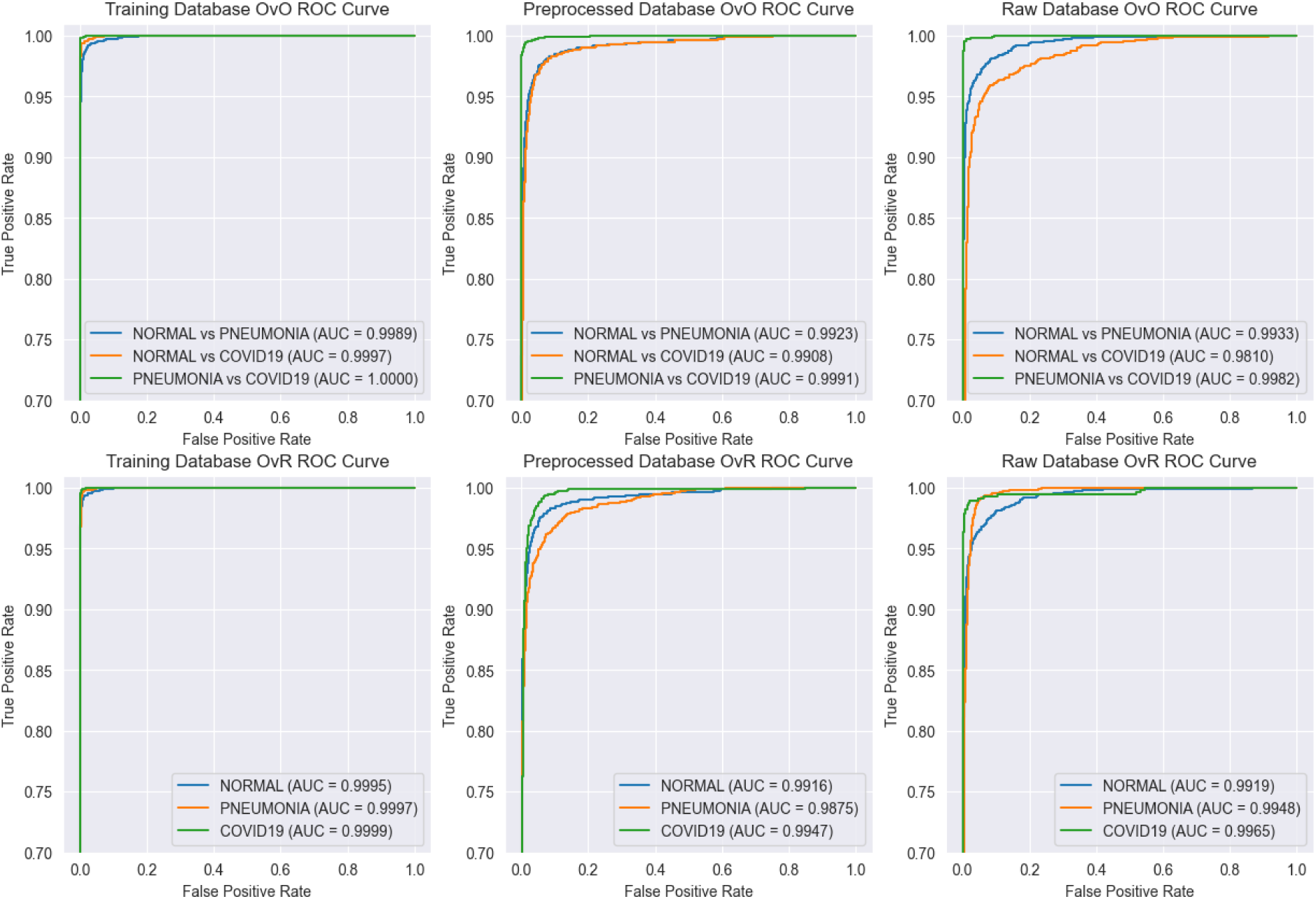
One-vs-One (OvO) and One-vs-Rest (OvR) Receiver Operating Characteristic (ROC) curves, and corresponding Area Under the Curve (AUC) scores for each database.

As can be seen in Figure 11, the proposed model shows superior AUC values for the COVID19 class, with overall the highest results in the databases for both the OvO and OvR approaches. This shows that the model classifies radiographs of patients with COVID-19 more reliably, which is the best possible result in terms of domain classes because a higher performance in diagnosing COVID 19 not only implies more safety and care for patients with this disease but also limits the possibility of contagion by overlooking a patient with SarsCov2. Overall, the model has excellent values for all AUC scores with a minimum value of 0.9810 and 0.9875 for the OvO and OvR approaches respectively.

In the context of the ROC Curve and AUC Score, it was found that there were no significant differences in the results obtained during the evaluation of the databases where the maximum difference in AUC score was 0.0083, observed between the training database with 0.9995 and the external preprocessed database with 0.9912.

The evaluation of the model on external databases yielded peculiar results. Contrary to expectations, the model performed best on the raw database, despite its previously described characteristics. Values for the accuracy metric obtained were 99.18%, 92.35%, and 96.78% for the training, preprocessed, and raw databases, respectively. These results are summarized in table 4 indicating a good generalization of the model, as it was able to achieve high accuracy on external databases with a large number of chest X-rays images. When evaluating external databases, the lowest accuracy result was observed on the preprocessed database, with a value of 92.35%. However, this result is particularly noteworthy as it exceeds the accuracy values reported on the test sets of several models previously studied in the literature [7, 8]. These findings demonstrate the robust generalization capabilities of the proposed approach on real-world data, which not only achieves high accuracy on previously unseen data, but also outperforms other models in the field.

**Table 4.**
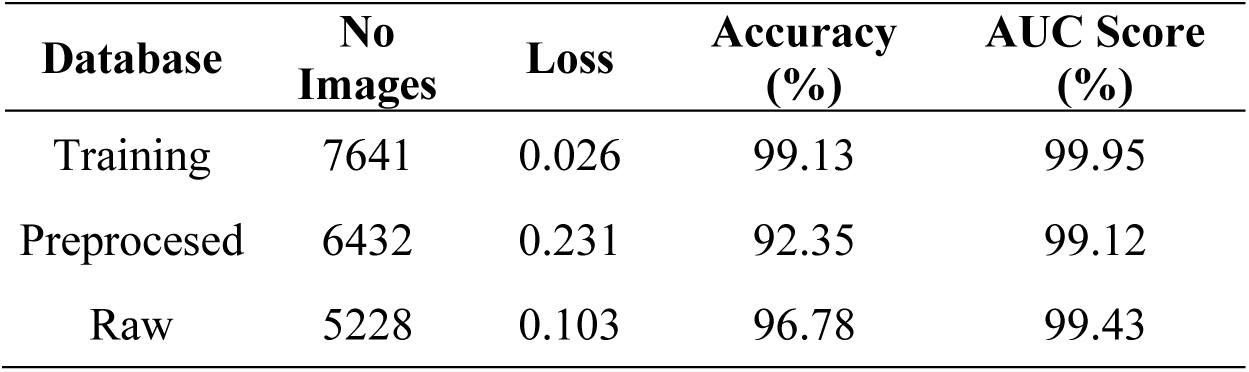
Metrics of the model evaluation on external databases.

A deeper evaluation of the model’s performance on external datasets was only performed on the preprocessed dataset, as it had the lowest performance during the model evaluation. The proposed X-COVNet model showed an accuracy of 92.35% on this dataset, misclassifying a total of 400 chest X-rays out of a total of 5228. The classification report shown in Table 5 details that the recall for COVID-19 showed the best value among the three classes, which is very close to 1. This means that the model missed very few patients with COVID-19 in the preprocessed dataset. On the other hand, a high precision of 98.97% for the Normal class indicates that the model is very accurate in predicting cases as normal. This means that there are few false positives for this class, which is crucial in a medical context as it reduces the likelihood of misdiagnosing a patient with a condition such as pneumonia or COVID-19 as Normal. These results are very reliable as the dataset is not biased towards any class and has a very balanced total for each class, such as 1802, 1800 and 1626 for the Normal, Pneumonia and COVID-19 classes respectively.

**Table 5.**
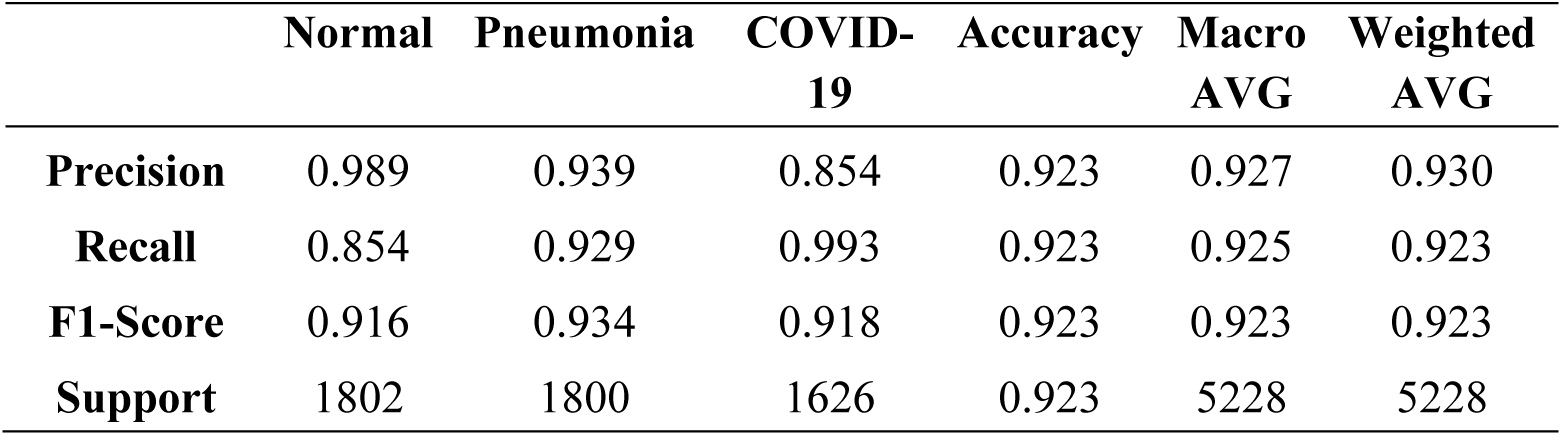
Classification Report of the External Preprocessed Dataset.

Table 6 below depicts the total number of misclassified images, categorized by each type of misclassification for further analysis. As mentioned above, the model rarely classifies patients with any disease, including pneumonia or COVID-19, as normal, with a total of 16 misclassifications. It is more likely that the model will classify a healthy patient as having a disease such as pneumonia or COVID-19, with a total of 263 incorrect predictions, or misclassify pneumonia as COVID-19, with a total of 120 misclassifications. However, it is highly unlikely that the model would miss a patient with COVID-19 with a total of 10 misclassifications in this evaluation. As mentioned in the class breakdown of the evaluation dataset, this is not due to any bias in the evaluation dataset. Thus, it supports the statement in the article that the model classifies patients with COVID-19 with high performance.

**Table 6.**
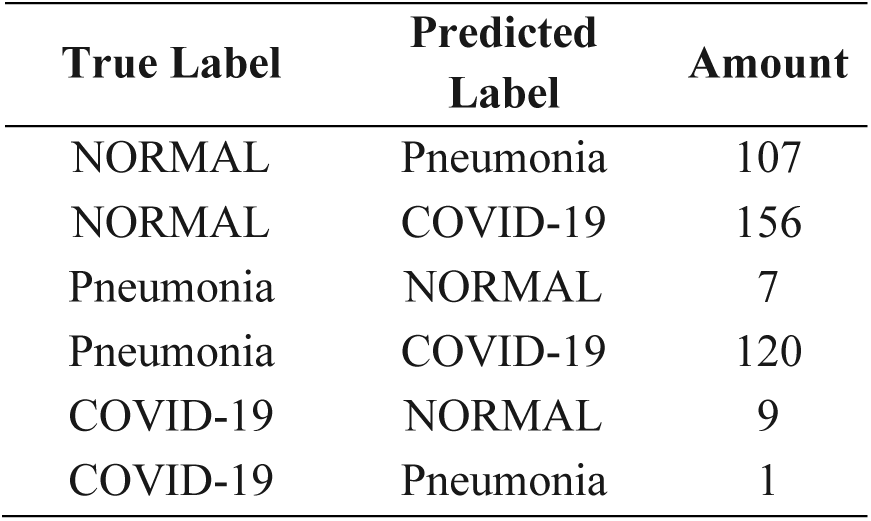
Distribution of total 400 misclassifications by category.

| True Label | Predicted Label | Amount |
| --- | --- | --- |
| NORMAL | Pneumonia | 107 |
| NORMAL | COVID-19 | 156 |
| Pneumonia | NORMAL | 7 |
| Pneumonia | COVID-19 | 120 |
| COVID-19 | NORMAL | 9 |
| COVID-19 | Pneumonia | 1 |

For visual understanding, Figure 12 shows a sample of misclassified images with the True Label (TL) plotted on the y-axis and the Predicted Label (PL) plotted on the x-axis.

**Fig. 12:**
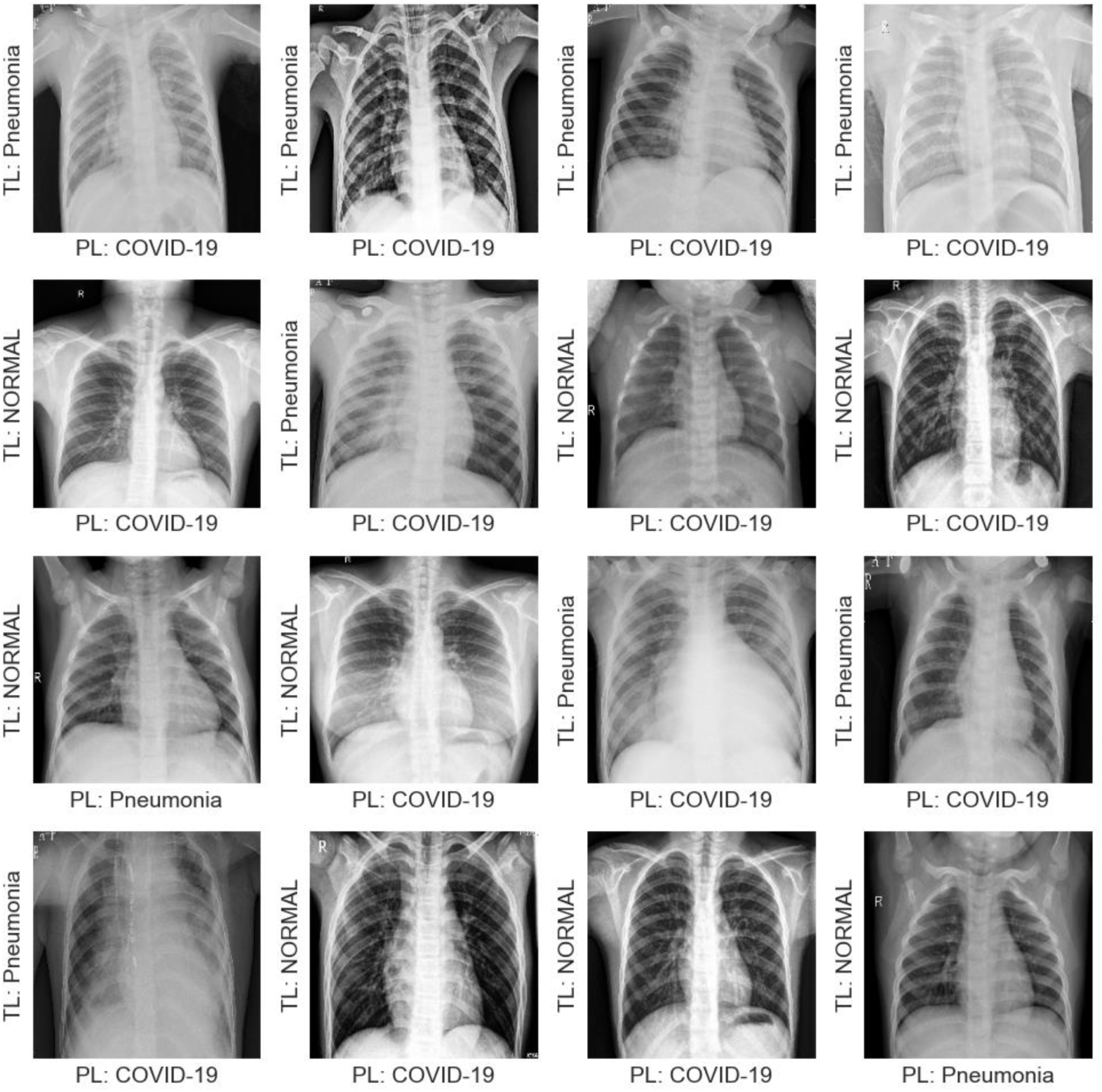
Random sample of misclassified images.

## 4. Discussion

### 4.1 Comparison with Published Models

During the comparison of the developed model with those reported in the literature, it was observed that, in contrast to the proposed model, 8 of the total 9 studied models for diagnosing chest radiographs rely on binary classifications. Results used during the comparison process were reported by their respective authors in their studies. The comparison of the models is carried out on the basis of the performance metrics obtained in the test set, as these are the ones most frequently collected in the research studied. While most of the literature reviewed utilized datasets with a magnitude of approximately 5000 images, the approach proposed in this study uses a larger dataset consisting of 7641 images.

Hashmi et al. [4] proposed a weighted classifier fed by 5 deep neural network architectures to classify normal or pneumonia in chest X-rays, achieving a recall value of 99.0%, higher than the 98.7% obtained by the proposed approach; however, their approach was slightly lower in terms of precision and accuracy metrics, with values of 98.3% and 98.4%, respectively. Ayan and Ünver [7] applied transfer learning and fine-tuning to the VGG16 and Xception architectures and obtained lower performance metrics than the proposed X-COVNet model, with accuracy, precision, and recall values of 87.0%, 87.0%, and 87.5%, respectively. The ensemble model proposed by Chouhan et al. [5] achieved recall and accuracy of 99.62% and 96.39%, respectively. Toğaçar et al. [6] achieved approximately 96.8% for recall, precision, and accuracy metrics. The approach proposed by Owida et al. [9] using the Mel Frequency Cepstral Coefficients method and a support vector machine classifier achieved 98.8% accuracy, unfortunately, others performance metrics were not reported in the research. The study conducted by Lee & Lim [10] achieved an accuracy of 99.90%, a precision of 98.99%, and a recall of 98.00%. Lee and Lim’s approach outperforms the proposed model only in terms of accuracy, with a difference of 0.84%. Jyoti et al. [11] achieved accuracies of 98.82% and 95.67% on small and large datasets respectively, which are less accurate than the proposed X-COVNet model. The approach proposed by Dalvi et al. [12], where data preprocessing is performed using the Nearest-Neighbors interpolation technique, achieved 96.37%, 94.08%, and 98.89% for accuracy, precision, and recall, respectively, which tends to be less performing than the proposed approach.

All of the models discussed above perform a binary classification of the chest radiograph, either for the COVID19/non-COVID19 or pneumonia/normal classes. In contrast to the aforementioned models, Civit-Masot et al. [8] used a training set of 316 images for a multiclass classification approach similar to the proposed approach, distinguishing between normal/healthy, pneumonia, and COVID-19 using the VGG16 architecture; they obtained 86.0% for each macro average of the metrics used in the comparison.

The model proposed in this paper generally showed better performance compared to the literature reviewed. The closest performance was achieved by Hashmi et al. [4], Lee & Lim [10], and Jyoti et al. [11], who used binary classification; however, the proposed approach outperformed these studies by using multi-class classification. Table 7 shows that the proposed X-COVNet model outperforms other approaches in general terms and demonstrates its superiority by achieving better metrics in a more complex classification task compared to most models in the reviewed literature.

**Table 7.** Comparison of the proposed model with published models.

| **Published Models** | **No. of Images** | **Precision (%)** | **Recall (%)** | **Accuracy (%)** | **Classification Type** | **External Validation** |
| --- | --- | --- | --- | --- | --- | --- |
| Civit-Masot et al., [8] | 316 | 86.00 | 86.00 | 86.00 | **Multi-class** | No |
| Ayan & Ünver [7] | 5856 | 87.00 | 87.50 | 87.00 | Binary | No |
| Chouhan et al., [5] | 5247 | 93.28 | **99.62** | 96.39 | Binary | No |
| Toğaçar et al., [6] | 5849 | 96.88 | 96.83 | 98.00 | Binary | No |
| Hashmi et al., [4] | 5856 | 98.26 | 99.00 | 98.43 | Binary | No |
| Owida et al., [9] | 5856 | - | - | 98.80 | Binary | No |
| Lee & Lim [10] | 5000 | 98.99 | 98.00 | **99.90** | Binary | No |
| Jyoti et al., [11] – Small | 2193 | **99.16** | 98.50 | 98.82 | Binary | No |
| Jyoti et al., [11] – Large | 5275 | 95.37 | 95.94 | 95.67 | Binary | No |
| Dalvi et al., [12] | 3010 | 94.08 | 98.89 | 96.37 | Binary | No |
| **Proposed Model (X-COVNet)** | **7641** | 98.99 | 98.73 | 99.06 | **Multi-class** | **Yes** |

The values are highlighted in bold to emphasize superior performance

### 4.2 Strengths and Limitations of the Proposed Model

A new computer-aided diagnostic model for chest X-ray has been achieved, capable of classifying patients with pneumonia-type lung disease. The model, which employs convolutional neural networks and transfer learning techniques, has demonstrated remarkable performance on validation and test data, achieving an accuracy of 0.9906 on test set. With its high accuracy and ability to differentiate between healthy patients, those with pneumonia and those with COVID-19, the model has the potential to improve the diagnosis of respiratory illnesses and patient care. The proposed model X-COVNet could serve as an adjunct to clinical decision making, it can assist radiologists in the decision-making process, but the final decision must be made by an expert. The proposed model is intended to support, not replace, the expertise of a trained radiologist in making a diagnosis [4].

The lack of external validation is a common problem in many research studies [4–12]. As explained earlier, external validation refers to the evaluation of a model’s performance on data not used during training or validation process. This is critical because it allows researchers to determine how well the model generalizes to real-world cases and can help identify potential biases or limitations in the model.

In the present research, an external validation was conducted and it was found that our proposed model showed high performance on external data. This is an important finding because it provides evidence that our model can accurately diagnose pneumonia-like lung diseases using chest X-rays, even when applied to data outside of our training dataset, which helps ensure that the model can accurately diagnose patients in a clinical setting.

It is also important to note that AI-based tools should be used with caution by trained professionals and not at the discretion of an individual. Misuse of these tools can pose risks to the health of patients and communities. Proper training and oversight are necessary to ensure that AI-based diagnostic tools are used safely and effectively in healthcare settings.

It should be acknowledged that the model has certain constraints. The Grad-CAM technique has not yet been implemented to facilitate the interpretation of the model’s classification decisions by medical specialists. However, work is actively being done to address this limitation and further improve the utility of the model for medical professionals.

In essence, this new model for computer-aided constitutes a substantial contribution in the field of computer-assisted diagnosis and has the capacity to profoundly influence both medical imaging and patient care. Through its ability to increase the accuracy and speed of diagnoses, reduce costs, and facilitate the use of remote medical consultations and monitoring, this model can greatly improve the accessibility, convenience, cost-effectiveness, and quality of healthcare for patients.

## 5. Conclusions and Future Work

The use of intelligent systems to improve the efficiency and effectiveness of medical diagnostics, as well as to reduce costs, is currently an accepted practice. In this research, a predictive model for computer-aided diagnosis of pneumonia-type pulmonary disease in chest X-rays based on Deep Convolutional Neural Networks using Transfer Learning through the Xception architecture was achieved with outstanding results and superior to those studied in the literature. The hyperparameter adjustment using Keras Tuner, the Learning Rate Reduction callback together with the image data augmentation and dropout layer allowed for improved the model’s performance. The several scores obtained throughout the model evaluation, accuracy, recall, precision and F1 score, demonstrated the reliability of the model.

Future research aims to integrate an algorithm for Gradient-weighted Class Activation Mapping (Grad-CAM) into the model to allow specialists understanding how the model has been driven to make the classification decision. Further, it is suggested to integrate the model with the help of CESIM into the Xavia-Pacs medical imaging tool deployed in Cuban medical centers.

## Author contributions

JF, JG, AO and DB contributed to conception and design of the study. JF and DB organized the databases. JF and DB performed the statistical analysis. JF and AO wrote the first draft of the manuscript. JF and AO wrote sections of the manuscript. All authors contributed to manuscript revision, read, and approved the submitted version.

## Competing interests

The author(s) declare(s) that there is no conflict of interest regarding the publication of this article.

## Data Availability

The X-COVNet model is publicly accessible and documented publicly on GitHub: https://github.com/WiseGeorge/X-COVNets. The repository aims to provide a library of open-source models for computer-aided diagnosis of pneumonia-like lung diseases.

## Notes

### Competing Interest Statement

The authors have declared no competing interest.

### Funding Statement

This study did not receive any funding

